# Dermal Phospho-α-Synuclein Among Individuals With and Without Cerebrospinal Fluid Aggregated α-Synuclein

**DOI:** 10.64898/2025.12.17.25342090

**Authors:** Lana M. Chahine, Krittika Chatterjee, Caroline Gochanour, David-Erick Lafontant, Rizwan S. Akhtar, Neil Shetty, Nabila Dahodwala, Sarah M. Brooker, Paulina Gonzalez-Latapi, Neha Prakash, Sara Defendorf, Jan Midyette, Tatiana Foroud, Kalpana M. Merchant, David G. Coughlin, Thomas F. Tropea, Mark Frasier, Danielle N. Larson, Christopher S. Coffey, Kenneth Marek, Tanya Simuni, the Parkinson’s Precision Medicine Initiative

## Abstract

**Background:** Dermal Serine-129-phosphorylated α-synuclein (dermal-pS129-αsyn) is a promising biomarker, but its accuracy for CNS aggregated α-synuclein (α-syn) is unknown. We examined sensitivity and specificity of dermal-pS129-αsyn for neuronal α-syn determined with CSF α-syn seed amplification assay (CSF α-syn SAA).

**Methods:** A sample of individuals enrolled in the Parkinson Precision Medicine Initiative (PPMI) with positive or negative CSF α-syn SAA underwent three 3-mm skin biopsies. Blinded assessment of dermal-pS129-αsyn was conducted. Participants were considered dermal-pS129-αsyn+ if ≥1 sections demonstrated evidence of ≥1 ps129-αsyn+ neuronal elements in any region biopsied.

**Results:** The sample consisted of 50 participants; PPMI enrollment cohort included PD (n=11), healthy control (n=5), and prodromal (RBD n=5, hyposmia n=27, non-manifesting carrier of pathogenic variant n=2). 30/38 CSF α-syn SAA+ were dermal-pS129-αsyn+; 6/12 CSF α-syn SAA-were dermal-pS129-αsyn-, yielding sensitivity and specificity of 79% and 50% respectively. 7/10 (70%) of participants with clinical diagnosis of PD who were CSF α-syn SAA+ were dermal-pS129-αsyn+. Of the 6 CSF α-syn SAA-cases who were dermal-pS129-αsyn+, 3 were enrolled in the HC cohort of PPMI, and 3 were prodromal (hyposmia) cases.

**Conclusion:** Low specificity for CSF α-syn SAA precludes use of dermal-pS129-αsyn as a marker of CNS neuronal α-syn aggregates. Future studies are needed to determine optimal methods to assess α-syn aggregates in central and peripheral compartments.

**Plain language summary:** Detecting abnormal alpha-synuclein in the skin may be a useful biomarker for Parkinson’s disease and other disorders where alpha-synuclein accumulates in brain cells. But whether skin alpha-synuclein reflects abnormal alpha-synuclein in brain cells is not known. Since alpha-synuclein in spinal fluid is an accurate reflection of alpha-synuclein in brain cells in almost all cases, we compared skin alpha-synuclein to alpha-synuclein in spinal fluid. We found that among people who have abnormal alpha-synuclein in spinal fluid, 79% have abnormal alpha-synuclein in skin. But among those who had negative (normal) alpha-synuclein in spinal fluid, 50% had abnormal skin alpha-synuclein. More studies are needed to understand what these results mean and the role of skin alpha-synuclein testing in Parkinson’s disease and other disorders where alpha-synuclein accumulates in brain cells.

## Introduction

Biomarkers of pathological α-synuclein (α-syn) that can be obtained with minimal invasiveness are needed for clinical research and in the clinic[1]. To this end, detection of abnormal α-syn in skin biopsies is promising[2]. A commonly used method involves double-immunostaining sections for neuronal elements and α-syn phosphorylated at Serine-129 residue. This co-localization is reported to improve specificity[3, 4]. Gibbons et al[5] reported a large, prospective multicenter study of dermal serine-129-phosphorylated α-synuclein (dermal-pS129-αsyn) detection by immunohistochemistry utilizing a commercially-available test (SynOne®). They demonstrated that 92.7% of clinically diagnosed PD and only 3.3% of controls were positive. There remains a gap in knowledge, however, as to the accuracy of dermal-pS129-αsyn for central nervous system (CNS) aggregated α-synuclein, for which the present gold standard is post-mortem neuropathological examination. A major recent advancement is availability of CSF α-syn seed amplification assay (CSF α-syn SAA), which has >95% sensitivity and specificity for autopsy-confirmed neocortical/limbic Lewy bodies[6]. Thus, CSF α-syn SAA serves as useful measure of CNS α-syn aggregates.

We conducted a pilot study to assess accuracy of dermal-pS129-αsyn for aggregated neuronal-predominant α-syn in CNS as measured *in vivo* with CSF α-syn SAA. Our objective was to assess sensitivity and specificity of dermal-pS129-αsyn for CSF α-syn SAA. Secondarily, we aimed to examine clinical characteristics of participants in relation to CSF and dermal-pS129-αsyn test results.

## Methods

This was a cross-sectional, observational multi-center study of a convenience sample of individuals enrolled in Parkinson Precision Medicine Initiative (PPMI) at 3 sites in the United States.

## Study Sample

PPMI inclusion/exclusion criteria have been published[7, 8]. Briefly, PPMI enrolls individuals with clinical diagnosis of PD, healthy controls (HC), non-manifesting carriers (NMC) of PD-associated pathogenic variants, or a prodromal group comprised of individuals with hyposmia, REM sleep behavior disorder (RBD), and/or biomarker abnormalities including positive CSF α-syn SAA and/or reduced dopamine transporter (DAT) SPECT binding[8].

This study aimed to recruit a convenience sample of 50 participants enrolled in PPMI. Inclusion criteria were CSF α-syn SAA+ among individuals enrolled in the PD cohort, CSF α-syn SAA-among individuals enrolled in HC cohort, and either CSF α-syn SAA– or CSF α-syn SAA+ among those enrolled in prodromal cohort. Exclusion criteria were type II seed CSF α-syn SAA (see below) or contraindications to skin biopsy.

Study activities were approved by Western-Copernicus Group Institutional Review Board. Informed consent was obtained from each participant.

Clinical Assessments, Imaging, and Genotyping

PPMI assessments of motor and non-motor symptoms of PD or dopaminergic imaging included in this analysis were:

– Movement Disorders Society Unified Parkinsons Disease Rating Scale (MDS-UPDRS)[9]
– REM sleep behavior disorder questionnaire [10]
– Scales for Outcomes in Parkinson’s-Autonomic (SCOPA-AUT)[11]
– Olfaction is assessed with the University of Pennsylvania Smell Identification Test-Revised (UPSIT-R), with normative values applied as described[12]
– Dopamine transporter (DAT) SPECT assessment was conducted as described, using the quantitative image analysis software MIAKAT (version 5.0) as described at ppmi-info.org [7, 13]
– Neuronal synuclein disease integrated staging system (NSD-ISS) stage was determined as described[14]

Genotyping: PD-associated pathogenic variants were determined where testing was available, as described at ppmi-info.org. Briefly, pathogenic variants may be identified in PPMI participants via one or more of the following methods: whole genome sequencing, whole exome sequencing, genome wide association studies, RNA-sequencing, Sanger sequencing of select GBA variants, or LRRK2 p.G2019S screening.

Cerebrospinal Fluid and Skin Biopsy Measures of α-syn

Participants underwent 3-mm skin biopsies from paracervical back, thigh, and distal leg per manufacturers’ protocol using SynOne® kit (CND Life Sciences, Scottsdale, AZ). Assessment of dermal-pS129-αsyn was performed as described[3] blinded to clinical features and CSF α-syn SAA status. For this analysis, participants were considered dermal-pS129-αsyn+ if ≥1 sections demonstrated evidence of ≥1 ps129-α-syn+ neuronal elements in any region biopsied.

Cerebrospinal fluid was tested at Amprion Diagnostics (San Diego, CA) as described[15, 16], using any one of 3 versions of the assay. Samples are designated as type I seed positive (CSF α-syn SAA+), inconclusive, negative (CSF α-syn SAA-), or (for the 24-hour reaction time assay) type II seed positive, as described[15–18].

When the study was initiated, mostly baseline visit CSF sample SAA results were available and were used to determine potential eligibility. For the primary analysis in this manuscript, cases who were type I seed CSF α-syn SAA+ at the closest available result up to the skin biopsy visit were considered CSF α-syn SAA+. Cases who were CSF α-syn SAA-at the closest available result up to skin biopsy visit were considered CSF α-syn SAA-. Where available, additional CSF samples were tested among CSF α-syn SAA-cases, and in select other cases.

## Statistical Analysis

Sample characteristics were summarized with descriptive statistics. Data from the visit at which skin biopsy was conducted were analyzed when available. Otherwise, for DAT and UPSIT, data acquired from the visit closest to skin biopsy (either before or after) were used. As mentioned, data for CSF α-syn SAA was acquired from the visit closest up until the time of skin biopsy.

Sensitivity and specificity of dermal-pS129-αsyn for presence or absence of type I seed CSF α-syn SAA was determined along with Wilson 95% confidence intervals (95%CIs).

In a sensitivity analysis, sensitivity and specificity were recalculated after excluding individuals who had CSF α-syn SAA results available only on CSF acquired >2 years before skin biopsy, or any case who had inconsistent or equivocal CSF results at any time point. Analyses were performed using SAS 9.4 (SAS/STAT 15.3; SAS Institute Inc., Cary, NC).

## Results

Of 50 participants in this study, 38 (76%) were type I seed CSF α-syn SAA+ and 12 (24%) were type I seed CSF α-syn SAA– (Table 1). Sample characteristics are summarized in Table 2 and characteristics of each participant are shown in Table 3. Two enrollments occurred (in error) in violation of the study protocol (i) 1 individual who was CSF α-syn SAA-was enrolled in the PD cohort; this participant was analyzed in the CSF α-syn SAA-group but was excluded in the sensitivity analysis, and (ii) 1 individual had a type II seed result in CSF when tested once but subsequently had negative CSF α-syn SAA (Supplementary Table 1). This individual was analyzed in the CSF α-syn SAA-group but was excluded in the sensitivity analysis.

**Table 1.** Sensitivity/Specificity of dermal phospho-α-syn for CSF α-syn seed amplification assay (SAA)

| | CSF α-syn SAA+ (N) | CSF α-syn SAA- (N) | Total N |
| --- | --- | --- | --- |
| **Dermal-ps129-α-syn+** | 30 | 6 | 36 |
| **Dermal-ps129-α-syn-** | 8 | 6 | 14 |
| **Total** | 38 | 12 | 50 |
| **Sensitivity** | 0.789 (95% CI<sup>†</sup> 0.637-0.889) |  |  |
| **Specificity** | 0.500 (95% CI<sup>†</sup> 0.254-0.746) |  |  |
| **Positive predictive value** | 0.833 (95% CI<sup>†</sup> 0.681-0.921) |  |  |
| **Negative predictive value** | 0.429 (95% CI<sup>†</sup> 0.214-0.674) |  |  |

<sup>†</sup>Wilson 95% confidence intervals (95%CI<sub>s</sub>)

**Table 2.** Demographic Characteristics by CSF α-syn SAA and skin biopsy result.

| Variable | Overall Results<br>(N = 50) | CSF α-syn SAA+ | | CSF α-syn SAA- | |
| --- | --- | --- | --- | --- | --- |
| | | Dermal-ps129-αsyn+<br>(N= 30) | Dermal-ps129-αsyn-<br>(N= 8) | Dermal-ps129-αsyn+<br>(N= 6) | Dermal-ps129-αsyn-<br>(N= 6) |
| **Sex, Male**, n (%) | 29 (58%) | 20 (67%) | 4 (50%) | 2 (33%) | 3 (50%) |
| **Age at skin biopsy (years)**, mean (SD) | 68.6 (6.1) | 68.6 (5.4) | 68.3 (4.5) | 62.7 (6.7) | 74.9 (5.8) |
| **Race, White**, n (%) | 49 (98%) | 30 (100%) | 7 (88%) | 6 (100%) | 6 (100%) |
| **Ethnicity, Hispanic/Latino**, n (%) | 1 (2%) | 1 (3%) | 0 | 0 | 0 |
| **Enrollment Cohort**, n (%) |  |  |  |  |  |
| PD* | 11 (22%) | 7 (23%) | 3 (38%) | 0 | 1 (17%) |
| Healthy Control | 5 (10%) | 0 | 0 | 3 (50%) | 2 (33%) |
| **Prodromal** |  |  |  |  |  |
| RBD** | 5 (10%) | 4 (13%) | 0 | 0 | 1 (17%) |
| Hyposmia Only*** | 27 (54%) | 18 (60%) | 5 (63%) | 3 (50%) | 1 (17%) |
| NMCs | 2 (4%) | 1 (3%) | 0 | 0 | 1 (17%) |
| **NSD Stage**, n (%) |  |  |  |  |  |
| N/A (Not NSD or Missing) | 12 | 0 | 0 | 6 | 6 |
| 1A | 1 (3%) | 1 (3%) | 0 | - | - |
| 1B | 1 (3%) | 1 (3%) | 0 | - | - |
| 2A | 8 (21%) | 8 (27%) | 0 | - | - |
| 2B | 9 (24%) | 7 (23%) | 2 (25%) | - | - |
| 3 | 10 (26%) | 8 (27%) | 2 (25%) | - | - |
| 4 | 8 (21%) | 5 (17%) | 3 (38%) | - | - |
| | | Dermal-ps129-αsyn+<br>(N= 30) | Dermal-ps129-αsyn-<br>(N= 8) | Dermal-ps129-αsyn+<br>(N= 6) | Dermal-ps129-αsyn-<br>(N= 6) |
| 5 | 1 (3%) | 0 | 1 (13%) | - | - |
| **MDS-UPDRS part I**, Median (IQR) | 5.5 (4.0, 10.0) | 5.5 (4.0, 10.0) | 10.5 (4.0, 17.5) | 3.5 (1.0, 6.0) | 6.0 (4.0, 7.0) |
| **MDS-UPDRS part II**, Median (IQR) | 3.0 (1.0, 5.0) | 3.0 (2.0, 5.0) | 5.5 (3.0, 14.0) | 0.5 (0.0, 2.0) | 2.0 (0.0, 3.0) |
| **MDS-UPDRS part III**, Median (IQR) | 4.0 (1.0, 14.0) | 5.0 (1.0, 14.0) | 6.0 (4.0, 31.0) | 1.5 (1.0, 3.0) | 1.0 (0.0, 1.0) |
| **MDS-UPDRS part total score**, Median (IQR) | 13.5 (7.0, 26.0) | 15.5 (8.5, 28.0) | 34.0 (16.0, 55.0) | 6.5 (3.0, 11.0) | 5.0 (4.0, 10.0) |
| **SCOPA-AUT total score**, Median (IQR) | 10.0 (7.0, 15.0) | 10.0 (8.0, 15.0) | 17.0 (10.5, 26.5) | 7.5 (3.0, 9.0) | 7.0 (0.0, 9.0) |
| **Latest Age/sex-expected low putamen ratio (MIAKAT)**, mean (SD) | 0.64 (0.29) | 0.59 (0.26) | 0.44 (0.23) | 0.84 (0.13) | 1.01 (0.16) |
| **Latest Revised UPSIT percentile $\leq 15$**, n (%) | 35 (70%) | 24 (80%) | 8 (100%) | 0 | 3 (50%) |
| **Time from LP to SB (years)**, Median (min, max) | 1.0 (0.0, 14.9) | 0.3 (0.0, 3.1) | 2.2 (0.2, 3.2) | 0.9 (0.0, 9.0) | 1.5 (1.0, 14.9) |
| **Time from UPSIT to SB (years)**, Median (min, max) | 0.3 (-0.9, 8.0) | 0.0 (0.0, 4.1) | 0.4 (-0.9, 4.0) | 1.5 (0.0, 4.8) | 3.4 (0.0, 8.0) |
| **Time from DAT to SB (years)**, Median (min, max) | 0.0 (-0.9, 14.9) | 0.0 (-0.0, 1.2) | 0.0 (-0.9, 3.3) | 1.6 (0.0, 14.0) | 0.6 (0.0, 14.9) |
| **Number of positive biopsies**, n (%) |  |  |  |  |  |
| Negative | 14 (28%) | 0 | 8 (100%) | 0 | 6 (100%) |
| One positive biopsy | 16 (32%) | 13 (43%) | 0 | 3 (50%) | 0 |
| Two positive biopsies | 15 (30%) | 12 (40%) | 0 | 3 (50%) | 0 |
| Three positive biopsies | 5 (10%) | 5 (17%) | 0 | 0 | 0 |
| **Only cervical region +**, n (%) | 10 (63%) | 8 (62%) | NA | 2 (67%) | NA |
| **Only thigh region +**, n (%) | 2 (13%) | 2 (15%) | NA | 0 | NA |
| **Only distal leg region +**, n (%) | 4 (25%) | 3 (23%) | NA | 1 (33%) | NA |
| | | Dermal-ps129-αsyn+<br>(N= 30) | Dermal-ps129-αsyn-<br>(N= 8) | Dermal-ps129-αsyn+<br>(N= 6) | Dermal-ps129-αsyn-<br>(N= 6) |
| **Evidence of small fiber neuropathy, n (%)** |  |  |  |  |  |
| Normal | 24 (48%) | 12 (40%) | 4 (50%) | 4 (67%) | 4 (67%) |
| Reduced | 25 (50%) | 17 (57%) | 4 (50%) | 2 (33%) | 2 (33%) |
| Other | 1 (2%) | 1 (3%) | 0 | 0 | 0 |
| **Cervical nerve fiber density, mean (SD)** | 31.6 (9.0) | 31.6 (9.0) | 28.5 (7.2) | 32.1 (10.0) | 35.0 (11.2) |
| **Thigh nerve fiber density, mean (SD)** | 11.8 (5.6) | 10.6 (2.2) | 12.7 (7.4) | 11.6 (2.5) | 17.0 (12.4) |
| **Distal leg nerve fiber density, mean (SD)** | 6.0 (2.6) | 5.6 (2.3) | 5.7 (3.0) | 7.8 (2.5) | 6.6 (3.1) |

All results are from the skin biopsy (SB) visit, except for CSF α-syn SAA, UPSIT, and DAT SPECT. CSF α-syn SAA results are based on the CSF sample collected up until SB. UPSIT and DAT SPECT results are from the assessment closest in time to SB, either before or after the SB visit.
Percentages for the 'only cervical,' 'only thigh,' and 'only distal leg' categories are calculated among participants with only 1 positive biopsy.
\* 1 PD cohort participant has pathogenic variant in GBA gene. \*\* 5 are PSG proven. \*\*\* 11 with DEB. The prodromal NMC group is comprised of 2 participants with pathogenic variants in known PD associated genes; specific variants are not shown for privacy reasons.
Ethnicity, Hispanic/Latino had 1 missing observation; MDS-UPDRS Part II and SCOPA-AUT total score each had 1 missing observation; MDS-UPDRS Part III had 3 missing observations; and MDS-UPDRS total score had 4 missing observations.
CSF α-syn SAA=Cerebrospinal fluid α-synuclein seed amplification assay; DAT=dopamine transporter SPECT; DEB=Dream enactment behavior; LP=lumbar puncture; MDS-UPDRS=Movement Disorders Society Modified Unified Parkinson's Disease Rating Scale; NMC=non-manifesting carrier (of parkinsonism-associated pathogenic variants); NSD-ISS=Neuronal Synuclein Disease Integrated Staging System; RBD=Rapid eye movement sleep behavior disorder; SB=skin biopsy; SCOPA-AUT=Scales for Outcomes in Parkinson's Disease - Autonomic Dysfunction; UPSIT=University of Pennsylvania Smell Identification Test.

**Table 3.** Clinical and biomarker characteristics of each participant enrolled in the study.

| *Enrollment Subgroup* | *NSD-ISS Stage* | *Sex* | *Age at SB (Y)* | *CSF α-syn SAA* | *Dermal p-α-syn* | *Cervical\** | *Thigh\** | *Distal Leg\** | *NFD* | *Age/sex-expected low putamen ratio* | *UPSIT-R percentile for age/sex* | *RBDS Q* | *SCOPA - AUT* | *LP to SB (y)\*\** | *DAT to SB (y)* | *UPSIT-R to SB (y)* |
| --- | --- | --- | --- | --- | --- | --- | --- | --- | --- | --- | --- | --- | --- | --- | --- | --- |
| Sporadic PD | 4 | Female | <60 | Positive | Positive | 2 | 0 | 0 | Present | 0.18 | 5 | 3 | 13 | 3.10 | 0.00 | 4.10 |
| Sporadic PD | 3 | Female | 60 - <=65 | Positive | Negative | 0 | 0 | 0 | Present | 0.23 | 10 | 7 | 11 | 2.97 | 0.00 | 3.95 |
| Sporadic PD | 3 | Male | 65 - <=70 | Positive | Positive | 0 | 0 | 2 | Absent | 0.42 | 14 | 0 | 2 | 2.18 | 0.00 | 2.18 |
| Sporadic PD | 3 | Female | <60 | Positive | Positive | 2 | 0 | 0 | Other | 0.29 | 3 | 2 | 2 | 0.00 | 0.00 | 2.10 |
| Sporadic PD | 5 | Male | 70 - <=75 | Positive | Negative | 0 | 0 | 0 | Absent | 0.24 | 5 | 1 | 25 | 3.16 | 3.28 | 3.16 |
| Sporadic PD | 3 | Male | 75 - <=80 | Positive | Positive | 0 | 2 | 1 | Absent | 0.53 | 15 | 2 | 18 | 2.97 | 1.21 | 2.97 |
| Sporadic PD | 3 | Male | 60 - <=65 | Positive | Negative | 0 | 0 | 0 | Present | 0.18 | 6 | 4 | 10 | 3.16 | -0.88 | 3.16 |
| Sporadic PD | 3 | Male | 65 - <=70 | Positive | Positive | 2 | 0 | 1 | Present | 0.32 | 2 | 0 | 8 | 0.00 | 0.00 | 2.28 |
| Sporadic PD | 3 | Female | <60 | Positive | Positive | 2 | 0 | 2 | Absent | 0.20 | 18 | 1 | 10 | 1.96 | 0.00 | 1.96 |
| Sporadic PD | 3 | Female | 65 - <=70 | Positive | Positive | 0 | 0 | 2 | Present | 0.26 | 16.5 | 3 | 5 | 1.94 | 0.00 | 1.94 |
| GBA PD | Not NSD | Male | >80 | Negative | Negative | 0 | 0 | 0 | Absent | 1.21 | 72.5 | 2 | 16 | 3.83 | 0.00 | 7.98 |
| RBD | 3 | Male | 70 - <=75 | Positive | Positive | 2 | 0 | 0 | Present | 0.38 | 23.5 | 10 | 19 | 0.00 | 0.00 | 0.00 |
| RBD | 4 | Female | 65 - <=70 | Positive | Positive | 0 | 2 | 2 | Present | 0.53 | 3 | . | . | 2.50 | 1.23 | 2.57 |
| RBD | 2b | Male | 65 - <=70 | Positive | Positive | 2 | 3 | 2 | Present | 0.41 | 15 | 12 | 4 | 0.00 | 0.00 | 0.00 |
| RBD | 2a | Male | 75 - <=80 | Positive | Positive | 0 | 3 | 2 | Absent | 0.76 | 3 | 12 | 14 | 2.27 | 0.00 | 0.00 |
| RBD | Not NSD | Male | 70 - <=75 | Negative | Negative | 0 | 0 | 0 | Absent | 0.99 | 99.5 | 11 | 0 | 1.02 | 0.00 | 0.00 |
| Hyposmia Only | Not NSD | Female | 65 - <=70 | Negative | Positive | 2 | 0 | 2 | Absent | 0.86 | 30 | 3 | 9 | 1.95 | 0.99 | 0.99 |
| Hyposmia Only | Not NSD | Female | 65 - <=70 | Negative | Positive | 0 | 0 | 2 | Present | 0.92 | 38.5 | 5 | 11 | 0.00 | 0.00 | 0.00 |
| Hyposmia Only | 2b | Female | 70 - <=75 | Positive | Negative | 0 | 0 | 0 | Absent | 0.62 | 1 | 4 | 16 | 1.13 | -0.91 | -0.90 |
| Hyposmia Only | 2a | Male | 75 - <=80 | Positive | Positive | 2 | 2 | 0 | Present | 0.85 | 8.5 | 2 | 11 | 0.00 | 0.00 | 0.00 |
| Hyposmia Only | 2b | Male | 60 - <=65 | Positive | Positive | 3 | 1 | 0 | Absent | 0.69 | 9 | 11 | 15 | 2.23 | 0.00 | 0.00 |
| Hyposmia Only | 2a | Male | 65 - <=70 | Positive | Positive | 2 | 1 | 2 | Present | 0.76 | 5 | 1 | 9 | 2.05 | 0.00 | 0.00 |
| Hyposmia Only | 1b | Male | 70 - <=75 | Positive | Positive | 0 | 2 | 2 | Present | 0.74 | 28 | 12 | 11 | 0.00 | 0.00 | 0.00 |
| Hyposmia Only | 2a | Male | 65 -<br>≤70 | Positive | Positive | 0 | 0 | 2 | Present | 0.99 | 3 | 1 | 5 | 0.00 | 0.00 | 0.00 |
| Hyposmia Only | 4 | Female | 65 -<br>≤70 | Positive | Negative | 0 | 0 | 0 | Absent | 0.64 | 3 | 6 | 28 | 2.22 | 0.00 | 0.00 |
| Hyposmia Only | 2b | Female | 70 -<br>≤75 | Positive | Positive | 0 | 2 | 0 | Present | 0.58 | 4 | 1 | 10 | 0.00 | 0.00 | 0.00 |
| Hyposmia Only | 1a | Female | 60 -<br>≤65 | Positive | Positive | 2 | 0 | 0 | Present | 0.77 | 73.5 | 2 | 9 | 0.00 | 0.00 | 0.00 |
| Hyposmia Only | 4 | Male | 75 -<br>≤80 | Positive | Negative | 0 | 0 | 0 | Absent | 0.74 | 1.5 | 6 | 30 | 2.12 | 0.00 | 0.00 |
| Hyposmia Only | 2b | Male | 75 -<br>≤80 | Positive | Positive | 2 | 0 | 0 | Present | 0.67 | 12.5 | 2 | 9 | 0.00 | 0.00 | 0.00 |
| Hyposmia Only | 4 | Male | 70 -<br>≤75 | Positive | Positive | 2 | 3 | 3 | Absent | 0.71 | 17.5 | 11 | 11 | 2.05 | 0.00 | 0.00 |
| Hyposmia Only | 3 | Male | 70 -<br>≤75 | Positive | Positive | 2 | 2 | 0 | Absent | 0.68 | 3 | 12 | 8 | 0.00 | 0.00 | 0.00 |
| Hyposmia Only | 2a | Male | 70 -<br>≤75 | Positive | Positive | 2 | 0 | 0 | Present | 1.00 | 1 | 4 | 7 | 1.13 | 0.00 | 0.00 |
| Hyposmia Only | 2b | Male | 65 -<br>≤70 | Positive | Negative | 0 | 0 | 0 | Present | 0.61 | 9.5 | 13 | 7 | 1.09 | 0.00 | 0.00 |
| Hyposmia Only | 2b | Male | 60 -<br>≤65 | Positive | Positive | 2 | 2 | 2 | Absent | 0.54 | 2 | 7 | 6 | 1.04 | 0.00 | 0.00 |
| Hyposmia Only | 4 | Male | 65 -<br>≤70 | Positive | Positive | 0 | 2 | 1 | Present | 0.32 | 9.5 | 10 | 25 | 1.05 | 0.00 | 0.00 |
| Hyposmia Only | Not NSD | Male | >80 | Negative | Negative | 0 | 0 | 0 | Present | 0.91 | 10.5 | 2 | 9 | 0.96 | 0.00 | 0.00 |
| Hyposmia Only | Not NSD | Female | 60 -<br>≤65 | Negative | Positive | 2 | 0 | 0 | Present | 0.62 | 31 | 3 | 8 | 0.75 | 0.00 | 0.00 |
| Hyposmia Only | 2b | Female | 65 -<br>≤70 | Positive | Positive | 2 | 0 | 2 | Absent | 0.38 | 1 | 9 | 8 | 0.29 | 0.00 | 0.75 |
| Hyposmia Only | 2b | Male | 70 -<br>≤75 | Positive | Positive | 0 | 2 | 0 | Absent | 0.68 | 13 | 9 | 16 | 0.25 | 0.00 | 0.70 |
| Hyposmia Only | 2a | Male | 65 -<br>≤70 | Positive | Positive | 1 | 1 | 0 | Absent | 1.08 | 15 | 3 | 10 | 0.20 | -0.04 | 0.52 |
| Hyposmia Only | 2a | Female | 65 -<br>≤70 | Positive | Positive | 2 | 0 | 0 | Present | 0.86 | 10 | 1 | 16 | 0.21 | 0.00 | 0.59 |
| Hyposmia Only | 2a | Male | 65 -<br>≤70 | Positive | Positive | 2 | 2 | 2 | Absent | 0.84 | 15 | 4 | 15 | 0.14 | 0.00 | 0.61 |
| Hyposmia Only | 4 | Female | 65 -<br>≤70 | Positive | Negative | 0 | 0 | 0 | Present | 0.24 | 4 | 8 | 18 | 0.23 | 0.00 | 0.71 |
| NMC | Not NSD | Female | 70 -<br>≤75 | Negative | Negative | 0 | 0 | 0 | Absent | 1.03 | 77 | 3 | 7 | 2.03 | 2.97 | 7.01 |
| NMC | 4 | Female | 60 -<br>≤65 | Positive | Positive | 2 | 0 | 0 | Present | 0.18 | 2 | 2 | 23 | 0.96 | 0.00 | 0.00 |
| Healthy Control | Not NSD | Male | 65 -<br>≤70 | Negative | Positive | 1 | 0 | 0 | Absent | 0.84 | 98.5 | 1 | 7 | 0.00 | 4.10 | 3.96 |
| Healthy Control | Not NSD | Male | 60 -<br>≤65 | Negative | Positive | 0 | 2 | 2 | Absent | 0.79 | 55.5 | 1 | 3 | 1.10 | 2.16 | 1.97 |
| Healthy Control | Not NSD | Female | 65 -<br>≤70 | Negative | Negative | 0 | 0 | 0 | Absent | 0.77 | 1 | 2 | 0 | 0.96 | 1.15 | 0.96 |
| Healthy Control | Not NSD | Female | 70 -<br><=75 | Negative | Negative | 0 | 0 | 0 | Present | 1.15 | 12 | 0 | 7 | 14.85 | 14.87 | 5.84 |
| Healthy Control | Not NSD | Female | <60 | Negative | Positive | 2 | 1 | 0 | Absent | 1.00 | 78.5 | 7 | 2 | 9.00 | 13.99 | 4.78 |

\* 0=No phosphorylated alpha-synuclein observed in stained sections. 1=1 colocalized fiber seen across all stained sections. 2=Two or more colocalized fibers seen across all stained sections. 3=At least one colocalized P-Syn positive nerve fiber observed in every stained section.
\*\*Time from CSF acquisition to skin biopsy, for the CSF sample for which the asyn α-syn SAA result was used for analysis.
CSF α-syn SAA=Cerebrospinal fluid alpha-synuclein seed amplification assay; DAT=dopamine transporter SPECT; HC=Healthy control; IU=Indiana University LP=lumbar puncture; MDS-UPDRS=Movement Disorders Society Modified Unified Parkinson's Disease Rating Scale; NFD=Nerve Fiber Density; NMC=non-manifesting carrier (of parkinsonism-associated pathogenic variants); NSD-ISS=Neuronal Synuclein Disease Integrated Staging System; RBD=Rapid eye movement sleep behavior disorder; SB=skin biopsy; SCOPA-AUT=Scales for Outcomes in Parkinson's Disease - Autonomic Dysfunction; UPSIT-R=University of Pennsylvania Smell Identification Test-Revised;

Among 38 CSF α-syn SAA+ cases, 30 were dermal-pS129-αsyn+; among 12 CSF α-syn SAA-cases, six were dermal-pS129-αsyn+, yielding sensitivity of 79% (95%CI: 64%-89%) and specificity of 50% (95%CI: 25%-75%).

Among the 12 CSF α-syn SAA-cases, DAT binding was >80^th^ percentile expected for age and sex in 5/6 prodromal cases, and olfaction was normal in 3/5 of HC. However, two HC participants had substantial intervals between CSF testing and skin biopsy (approximately 15 and 9 years, respectively); the corresponding intervals between DAT SPECT and skin biopsy were approximately 15 and 14 years, and between olfactory testing and skin biopsy were approximately 6 and 5 years, respectively. Recalculation of sensitivity and specificity in n=32 participants (restricting to those who had CSF testing ≤2 years up to skin biopsy and excluding two enrollments that did not meet inclusion/exclusion criteria, indicated higher sensitivity (88%; 95%CI: 70%-96%) but lower specificity (43%; 95%CI: 16%-75%) (Supplementary Table 2). For the 3 HC who were CSF α-syn SAA– and dermal-pS129-αsyn+, additional CSF samples acquired closer to skin biopsy were tested and results demonstrated a persistent CSF α-syn SAA-result (Supplementary Table 1).

Dermal-ps129-α-syn and CSF α-syn SAA results according to enrollment cohort/clinical diagnosis are shown in Table 4. Among the type I seed CSF α-syn SAA+ cases, 7/10 (70%) sporadic PD, 4/4 (100%) RBD, 18/23 (78%) with hyposmia, and 1 non-manifesting carrier of PD-related pathogenic variant were dermal-pS129-αsyn+. As for CSF α-syn SAA+ dermal-pS129-αsyn-cases, 3 were in the sporadic PD group and 5 were enrolled in the prodromal cohort based on presence of hyposmia. Among the hyposmia group, dermal-pS129-αsyn result was consistent with CSF result in 19/27 cases. Dermal-ps129-α-syn in the hyposmia group did not vary according to whether the participant had possible RBD (pRBD) symptoms.

**Table 4.**
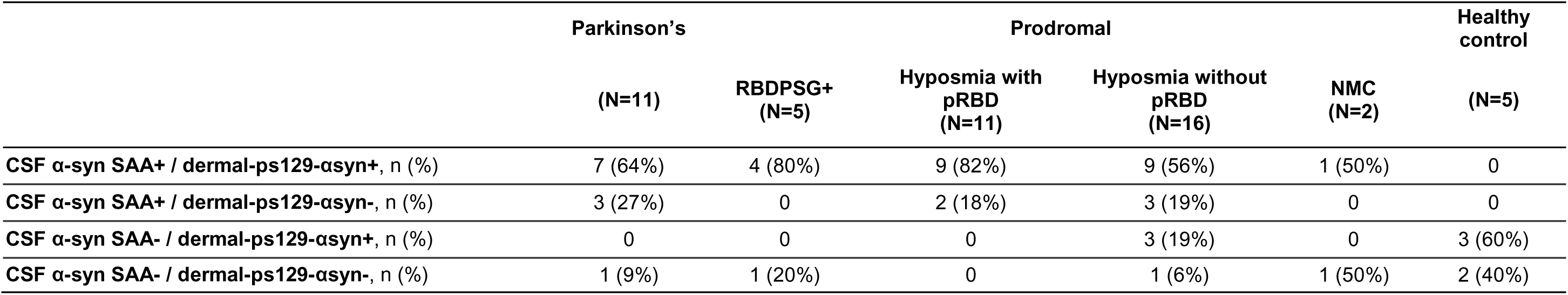
CSF α-syn SAA and dermal results according to enrollment cohort (clinical diagnosis).

|  | Parkinson's |  | Prodromal |  |  | Healthy control |
| --- | --- | --- | --- | --- | --- | --- |
|  | (N=11) | RBDPSG+<br>(N=5) | Hyposmia with<br>pRBD<br>(N=11) | Hyposmia without<br>pRBD<br>(N=16) | NMC<br>(N=2) | (N=5) |
| CSF α-syn SAA+ / dermal-ps129-αsyn+, n (%) | 7 (64%) | 4 (80%) | 9 (82%) | 9 (56%) | 1 (50%) | 0 |
| CSF α-syn SAA+ / dermal-ps129-αsyn-, n (%) | 3 (27%) | 0 | 2 (18%) | 3 (19%) | 0 | 0 |
| CSF α-syn SAA- / dermal-ps129-αsyn+, n (%) | 0 | 0 | 0 | 3 (19%) | 0 | 3 (60%) |
| CSF α-syn SAA- / dermal-ps129-αsyn-, n (%) | 1 (9%) | 1 (20%) | 0 | 1 (6%) | 1 (50%) | 2 (40%) |

For privacy reasons, variant information for the 2 non-manifesting carriers of PD-associated variants included in this study are not provided here. As for other genetic test results, in 3 cases, all in the CSF α-syn SAA-group, GBA 409S variant was identified. Of note, 1 was enrolled in the HC group and was dermal-pS129-αsyn+. 33 cases had whole genome sequencing data available for this analysis and did not identify pathogenic variants in PD-related genes. Seven participants did not have WGS available but had been tested for LRRK2 G2019S variant and were negative. Genetic data were unavailable for inclusion in this analysis for the remainder of participants.

## Discussion

With the advent of *in vivo* biomarkers of α-syn, biologic definition of synucleinopathies is now possible[19, 20]. Biologic definition of disease has important applications in clinical trials, where biomarker-based detection of abnormal α-syn may be used as an inclusion criterion or to stratify individuals when analyzing response to therapeutics. The CSF α-syn SAA used in this study has undergone rigorous technical and clinical validation against postmortem findings and is established as a highly accurate biomarker of neocortical/limbic neuronal aggregated α-syn. A relative barrier to widespread use of this test is the requirement to obtain CSF, and a more easily obtained test that accurately reflects CNS α-syn pathology is crucial. To that end, and given promising data on dermal-pS129-αsyn, we examined sensitivity and specificity of dermal-pS129-αsyn for CSF α-syn SAA. We found that dermal-pS129-αsyn has a moderate sensitivity of 79% for type I seed CSF α-syn SAA+. However, specificity of dermal-pS129-αsyn for CSF α-syn SAA-was low at 50%.

Sensitivity in our sample, whether for CSF α-syn SAA or for clinical diagnosis of PD, is consistent with the majority of published data; dermal p-α-syn positivity has been reported in 70%-100% of PD cases[2]. This variability has been attributed to clinical misdiagnosis and methodological differences (specimen processing, antibody used, slice thickness, detection methods (immunohistochemistry vs immunofluorescence), number of biopsies etc). In addition, it is possible that in some individuals with central neuronal synucleinopathy there is minimal peripheral synucleinopathy; studies that include CSF aSynSAA, dermal-pS129-αsyn testing, and/or autopsy are limited so these relationships are not fully known. Our data does not indicate that small fiber neuropathy is a likely explanation. It is also possible that the dermal-pS129-αsyn-cases are false negative, whether due to the patchy nature of peripheral α-syn or that the p129-α-syn antibody may not detect some forms of pathologic α-syn such as those truncated at or before the Serine129 residue[21].

When interpreting the low specificity of dermal-pS129-αsyn for CSF α-syn SAA in our study, there are a few possibilities. One is a false negative CSF result. Cerebrospinal fluid α-syn SAA is positive in >95% of cases with neocortical Lewy body pathology on autopsy. However, a small proportion of individuals who are CSF α-syn SAA-may in fact have CNS α-syn pathology. Some may have aggregated α-syn that is not neuronal-predominant. Others may be false negatives; this is more likely among cases with focal Lewy pathology[6]. The second possibility is that dermal-pS129-αsyn testing with the utilized method may be detecting non-pathologic α-syn. Indeed, while p129-α-synuclein concentration is highest in aggregated α-syn, under physiologic conditions a fraction of normal α-syn may also be phosphorylated at this site[22]. Future studies need to investigate these possibilities rigorously. The third explanation is that dermal-pS129-αsyn is detecting pathologic α-syn that is present only peripherally. The occurrence of this phase of synucleinopathy has been hypothesized in the “body-first” versus “brain-first” model of Lewy Body Disorders, which proposes that in some individuals, initial pathogenic α-synuclein originates peripherally followed by spread to the CNS[23]. Importantly, autopsy and/or *in vivo* data supporting the occurrence of periphery-*first* synucleinopathy are quite limited—few data demonstrate presence of abnormal α-syn peripherally in absence of abnormal α-syn centrally. However, assuming that such a phase exists, presence of dermal-pS129-αsyn in absence of CSF α-syn SAA could theoretically be a reflection of it. In this context, the time between CSF and skin biopsy acquisition could theoretically be relevant. Results of testing of CSF acquired longitudinally in a subset of our sample, as well as in the PPMI cohort at large in a separate study[24] indicate high intra-individual consistency of CSF aSyn SAA results over time. However, longitudinal studies, with repeat sampling of both skin, CSF, and DAT SPECT, will be needed to investigate these questions systematically.

Two other studies examined dermal-pS129-αsyn in relation to CSF α-syn SAA[25, 26]. Liguori et al [25] assessed dermal-pS129-αsyn and CSF α-syn SAA in cases with iRBD, other sleep disorders or neuropathy. Sensitivity and specificity of dermal-pS129-αsyn for CSF α-syn SAA was 58% and 75% respectively. In another study by Donadio et al[26], 6/6 CSF α-syn SAA+ cases were dermal p-α-syn-129+. All but 1 of the 28 CSF α-syn SAA-case was dermal-pS129-αsyn-yielding 100% sensitivity and 96% specificity. Comparing our results to these studies is made challenging by differences in methods for CSF α-syn SAA and dermal-pS129-αsyn detection[27].

Limitations of this study include the small sample size, cross sectional study design, lack of racial/ethnic diversity of the sample, and the prolonged time between assessments in a subset of participants. However, while at the time of skin biopsy there was a prolonged time between CSF acquisition and skin biopsy for a few participants, additional CSF was acquired and CSF α-syn SAA results at later time points (closer to skin biopsy) remained consistent.

Minimally invasive biomarkers, such as accurate skin α-synuclein measures, are critically needed in the research setting and in clinic. A very high specificity for dermal-pS129-αsyn for clinically-defined diagnosis in both early parkinsonism with 3-4 years’ follow-up[28], as well as in more advanced PD [5] have been reported. In contrast, our data indicate that dermal-pS129-αsyn is not specific for CNS aggregated α-syn as determined with CSF α-syn SAA. Our results do not necessarily negate the value of dermal-pS129-αsyn testing in individuals with parkinsonism and those at risk for it. Rather, they highlight the need for additional data to help interpret results of central versus peripheral biomarkers of α-syn pathology and their relationship to each other. Specifically, studies by multiple independent groups are needed that collect both CSF and skin biopsy obtained concurrently, as well as thorough application of clinical diagnostic criteria with longitudinal follow-up.

## Data Availability

Data availability: All data were obtained on 15 June 2026 from the PPMI database (www.ppmi-info.org/access-data-specimens/download-data), RRID:SCR_006431. Analyses, conducted by PPMI Statistics Core, used actual dates, a restricted (not publicly available) data element. PPMI Data Access Committee approved use of CSF α-syn SAA and DaTscan results. Statistical analysis codes are shared on Zenodo [10.5281/zenodo.17645460]. PPMI protocol information is available at: https://dx.doi.org/10.17504/protocols.io.n92ldmw6ol5b/v2.

## Acknowledgments

The authors thank the PPMI study participants.

PPMI – a public-private partnership – is funded by the Michael J. Fox Foundation for Parkinson’s Research and funding partners, including AbbVie, Alamar Biosciences, Aligning Science Across Parkinson’s (ASAP), Arrowhead Pharma, Arvinas, AskBio, BIAL, BioArctic, Biohaven, BlueRock Therapeutics, Bristol Myers Squibb, Calico Labs, Capsida Biotherapeutics, Critical Path Institute, DaCapo Brainscience, Denali, Edmond J. Safra Foundation, Eli Lilly, Gain Therapeutics, GE Healthcare, Genentech, GSK, Insitro, Johnson & Johnson Innovative Medicine, Lundbeck, Merck, Neumora, Neuron23, Novartis, Olink, Regeneron, Roche, Sanofi, Tenvie, UCB, Vanqua Bio, Voyager Therapeutics, The Weston Family Foundation.

## Author contributions

Design: Lana M Chahine, Chris Coffey, Ken Marek, Tanya Simuni

Execution: Lana M Chahine, Krittika Chatterjee, Caroline Gochanour, David-Erick Lafontant, Rizwan Akhtar, Neil Shetty, Nabila Dahodwala, Sarah Brooker, Paulina Gonzalez-Latapi, Neha Prakash, Sara Defendorf, Jan Midyette, Kalpana Merchant, David Coughlin, Thomas F Tropea, Mark Frasier, Chris Coffey, Ken Marek, Tanya Simuni

Analysis: Krittika Chatterjee, Caroline Gochanour, David-Erick Lafontant, Chris Coffey, Lana Chahine

Writing: Lana M Chahine, Tanya Simuni

Editing of final version of the manuscript: Lana M Chahine, Krittika Chatterjee, Caroline Gochanour, David-Erick Lafontant, Rizwan Akhtar, Neil Shetty, Nabila Dahodwala, Sarah Brooker, Paulina Gonzalez-Latapi, Neha Prakash, Sara Defendorf, Jan Midyette,Tatiana Foroud, Kalpana Merchant, David Coughlin, Thomas F Tropea, Mark Frasier, Chris Coffey, Ken Marek, Tanya Simuni

## Statements and Declarations

Ethical considerations: Study activities were approved by Western-Copernicus Group Institutional Review Board.

Consent to participate: Informed consent was obtained from each participant.

Consent for publication: not applicable

Declaration of conflicting interest: The author(s) declared no potential conflicts of interest with respect to the research, authorship, and/or publication of this article’.

Funding statement: This study was funded by The Michael J. Fox Foundation for Parkinson’s Research, Grant ID: 026366/026110.

