## Supplement Materials for "Dermal Phospho-α-Synuclein Among Individuals With and Without Cerebrospinal Fluid Aggregated α-Synuclein"

**Supplementary Table 1.** Longitudinal CSF α-Synuclein Seed Amplification Assay Results

| **Enrollment Subgroup** | **SB result** | **PPMI BL to SB (y)** | **LP at SB (y)*** | **LP at SB Result** | **LP1 to SB (y)** | **Result1** | **LP2 to SB (y)** | **Result2** | **LP3 to SB (y)** | **Result3** |
| --- | --- | --- | --- | --- | --- | --- | --- | --- | --- | --- |
| Sporadic PD | Positive | 4.10 | 3.10 | Positive | 4.10 | Positive | 3.10 | Positive |  |  |
| Sporadic PD | Negative | 3.95 | 2.97 | Positive | 3.93 | Positive | 2.97 | Positive |  |  |
| Sporadic PD | Positive | 2.18 | 2.18 | Positive | 2.18 | Positive |  |  |  |  |
| Sporadic PD | Positive | 2.10 | 0.00 | Positive | 2.10 | Positive | 0.00 | Positive |  |  |
| Sporadic PD | Negative | 3.16 | 3.16 | Positive | 3.16 | Positive |  |  |  |  |
| Sporadic PD | Positive | 2.97 | 2.97 | Positive | 2.97 | Positive |  |  |  |  |
| Sporadic PD | Negative | 3.16 | 3.16 | Positive | 3.16 | Positive |  |  |  |  |
| Sporadic PD | Positive | 2.28 | 0.00 | Positive | 2.28 | Positive | 0.00 | Positive |  |  |
| Sporadic PD | Positive | 1.99 | 1.96 | Positive | 1.96 | Positive |  |  |  |  |
| Sporadic PD | Positive | 1.94 | 1.94 | Positive | 1.94 | Positive |  |  |  |  |
| GBA PD | Negative | 7.98 | 3.83 | Negative | 7.98 | Negative | 3.83 | Negative |  |  |
| RBD | Positive | 2.04 | 0.00 | Positive | 2.04 | Positive | 0.00 | Positive |  |  |
| RBD | Positive | 2.50 | 2.50 | Positive | 2.50 | Positive |  |  |  |  |
| RBD | Positive | 2.15 | 0.00 | Positive | 2.15 | Positive | 0.00 | Positive |  |  |
| RBD | Positive | 2.27 | 2.27 | Positive | 2.27 | Positive |  |  |  |  |
| RBD | Negative | 1.02 | 1.02 | Negative | 1.02 | Negative |  |  |  |  |
| Hyposmia Only | Positive | 2.98 | 1.95 | Negative | 2.96 | Negative | 1.95 | Negative |  |  |
| Hyposmia Only | Positive | 2.09 | 0.00 | Negative | 2.08 | Type II Positive | 1.00 | Negative | 0.00 | Negative |
| Hyposmia Only | Negative | 3.18 | 1.13 | Positive | 3.18 | Positive | 1.13 | Positive |  |  |
| Hyposmia Only | Positive | 2.18 | 0.00 | Positive | 2.18 | Positive | 0.00 | Positive |  |  |
| Hyposmia Only | Positive | 2.23 | 2.23 | Positive | 2.23 | Positive |  |  |  |  |
| Hyposmia Only | Positive | 2.15 | 2.05 | Positive | 2.05 | Positive |  |  |  |  |
| Hyposmia Only | Positive | 2.03 | 0.00 | Positive | 2.01 | Positive | 0.00 | Positive |  |  |
| Hyposmia Only | Positive | 2.07 | 0.00 | Positive | 2.07 | Positive | 0.00 | Positive |  |  |
| Hyposmia Only | Negative | 2.22 | 2.22 | Positive | 2.22 | Positive |  |  |  |  |
| Hyposmia Only | Positive | 2.03 | 0.00 | Positive | 2.03 | Positive | 0.00 | Positive |  |  |
| Hyposmia Only | Positive | 2.05 | 0.00 | Positive | 2.05 | Positive | 0.00 | Positive |  |  |
| Hyposmia Only | Negative | 2.12 | 2.12 | Positive | 2.12 | Positive |  |  |  |  |
| Hyposmia Only | Positive | 2.22 | 0.00 | Positive | 2.22 | Positive | 0.00 | Positive |  |  |
| Hyposmia Only | Positive | 2.17 | 2.05 | Positive | 2.05 | Positive |  |  |  |  |
| Hyposmia Only | Positive | 2.09 | 0.00 | Positive | 2.09 | Positive | 0.00 | Positive |  |  |
| Hyposmia Only | Positive | 1.13 | 1.13 | Positive | 1.13 | Positive | -1.04 | Positive |  |  |
| Hyposmia Only | Negative | 1.09 | 1.09 | Positive | 1.09 | Positive | -0.94 | Positive |  |  |
| Hyposmia Only | Positive | 1.05 | 1.04 | Positive | 1.04 | Positive | -0.87 | Positive |  |  |
| Hyposmia Only | Positive | 1.05 | 1.05 | Positive | 1.05 | Positive | -1.00 | Positive |  |  |
| Hyposmia Only | Negative | 0.96 | 0.96 | Negative | 0.96 | Negative |  |  |  |  |
| Hyposmia Only | Positive | 1.02 | 0.75 | Negative | 0.75 | Negative |  |  |  |  |
| Hyposmia Only | Positive | 0.00 | 0.29 | Positive | 0.29 | Positive |  |  |  |  |
| Hyposmia Only | Positive | 0.00 | 0.25 | Positive | 0.25 | Positive |  |  |  |  |
| Hyposmia Only | Positive | 0.00 | 0.20 | Positive | 0.20 | Positive |  |  |  |  |
| Hyposmia Only | Positive | 0.00 | 0.21 | Positive | 0.21 | Positive |  |  |  |  |
| Hyposmia Only | Positive | 0.00 | 0.14 | Positive | 0.14 | Positive |  |  |  |  |
| Hyposmia Only | Negative | 0.00 | 0.23 | Positive | 0.23 | Positive |  |  |  |  |
| NMC | Negative | 7.01 | 2.03 | Negative | 7.01 | Negative | 2.03 | Negative |  |  |
| NMC | Positive | 0.96 | 0.96 | Positive | 0.96 | Positive | -0.96 | Positive |  |  |
| Healthy Control | Positive | 3.97 | 0.00 | Negative | 3.96 | Negative | 0.99 | Negative | 0.00 | Negative |
| Healthy Control | Positive | 1.97 | 1.10 | Negative | 1.96 | Negative | 1.10 | Negative | -1.25 | Negative |
| Healthy Control | Negative | 0.96 | 0.96 | Negative | 0.96 | Negative |  |  |  |  |
| Healthy Control | Negative | 14.83 | 14.85 | Negative | 14.85 | Negative |  |  |  |  |
| Healthy Control | Positive | 13.95 | 9.00 | Negative | 13.95 | Negative | 9.00 | Negative | -1.14 | Negative |

LPs are numbered in the order of their occurrence. When LP to SB years = 0, this signifies the result of the CSF alpha-syn SAA when CSF was collected at the same visit as the skin biopsy.

*This column indicates the LP closest to skin biopsy visit, up to and including the time of skin biopsy, and the CSF result used in the main analysis. For each participant, this is equivalent to either the LP1, LP2, or LP3 columns as specified.

**Supplementary Table 2.** Sensitivity/Specificity of dermal phospho-α-syn for CSF α-syn SAA when including participants with CSF results tested within 2 years of skin biopsy and excluding 2 participants who did not meet enrollment criteria (1 participant who was CSF α-syn SAA- who was enrolled in the PD cohort and 1 individual who had a type II CSF result in CSF when tested once (but subsequently had negative CSF α-syn SAA))

|  | **CSF α-syn SAA+ (N)** | **CSF α-syn SAA- (N)** | **Total N** |
| --- | --- | --- | --- |
| **Dermal-ps129-α-syn+** | 22 | **4** | 26 |
| **Dermal-ps129-α-syn-** | 3 | 3 | 6 |
| **Total** | 25 | 7 | 32 |
| **Sensitivity** | 0.880 (95% CI 0.700-0.958) | | |
| **Specificity** | 0.429 (95% CI 0.158-0.750) | | |
| **Positive predictive value** | 0.846 (95% CI 0.665-0.938) | | |
| **Negative predictive value** | 0.500 (95% CI 0.188-0.812) | | |
